# Phage Therapy: Navigating the Mechanisms, Benefits, and Challenges in the Fight Against Multidrug-Resistant Infections

**DOI:** 10.1101/2025.04.19.25325436

**Authors:** Mohammed Munshi, Sabrina Jahangir, Sumaiya Sumaiya

**Author notes:** Corresponding Author: Name: Mohammed Munshi, Postal Address: House-6, Lane-21, Block-C, Avenue-5, Mirpur-11Pallabi, Dhaka-1216, Bangladesh, Contact No.

## Abstract

**Background:** The emergence of multidrug-resistant (MDR) infections represents one of the most critical challenges faced by contemporary medical practice. The World Health Organization (WHO) has identified antibiotic resistance as a significant threat to global health, with MDR bacteria leading to increased morbidity, mortality, and healthcare costs ^[1]^.

**Objective (PICOS):** This review examines ***(P)*** MDR infection patients receiving ***(I)*** short-term (0-14 day) phage therapy versus ***(C)*** long-term/antibiotic treatments, assessing ***(O)*** efficacy (eradication rates) and safety (adverse events) through ***(S)*** clinical trial/case study analysis.

**Primary & Secondary Outcome Measures:** Efficacy (bacterial eradication) and safety (adverse events) of 0–14 day phage therapy, compared to conventional antibiotics or longer phage regimens.

**Intervention:** Application of bacteriophages (viruses that infect and lyse bacteria) as targeted antimicrobial agents.

**Methods:** A comprehensive review of clinical trials, case studies, and experimental models from peer-reviewed literature.

**Results:** Phage therapy demonstrates high bacterial specificity, adaptability to resistance, and synergistic effects with antibiotics.

**Article Summary:** While phage therapy offers a promising alternative to antibiotics, its clinical integration faces regulatory, logistical, and safety challenges.

**Strengths and limitations of this study:** The strengths include detailed analysis of phage mechanisms, clinical applications, and therapeutic potential. The limitations are limited large-scale randomized controlled trials (RCTs) and standardized treatment protocols & last is This review was not registered in PROSPERO.

**Conclusion:** Phage therapy holds transformative potential in combating MDR infections but requires further research, regulatory standardization, and clinical validation.

## INTRODUCTION

The global medical emergency presents itself as multidrug-resistant (MDR) infections because standard antibiotics lose their effectiveness in treating these pathogens. The World Health Organization ranks antibiotic resistance as one of the ten major threats to public health because **10 million annual deaths will occur by 2050** if resistance is unchecked (WHO, 2024) ^[1]^. The medical approach of bacteriophage therapy (also known as phage therapy) has re-emerged as an effective antibiotic replacement for modern medicine in this specific environment ^[2]^. This review follows PRISMA 2020 guidelines.

### The Crisis of Antibiotic Resistance

Medicinal approaches known as traditional antibiotics have gradually become less effective for several reasons:

- Overprescription and misuse in healthcare and agriculture ^[3]^.
- Horizontal gene transfer among bacteria, accelerating resistance spread ^[4]^.
- Limited development of novel antibiotics due to pharmaceutical industry disinvestment ^[5]^.

### Phage Therapy: A Resurgent Solution

Phage therapy, developed as a treatment approach in the first part of the 20th century, provides distinctive benefits to medicine.

1. The specificity of phages remains high because they exclusively attack disease-causing bacteria without harming beneficial microorganisms in the body ^[6]^.
2. At the infection site phages continue their replication to increase their therapeutic power ^[7]^.
3. Phages demonstrate the ability to adapt their structures when bacteria evolve resistance so they can overcome these mechanisms ^[8]^.

Opposing widespread adoption of phage therapy are the regulatory barriers, together with bacterial resistance to phages and concerns about safety ^[9]^.

### Research Significance

The investigation sought to determine if phage therapy holds promise as a treatment option for multidrug-resistant (MDR) infections through evaluation of its operational modes and clinical effectiveness as well as its facing obstructions. The study examined the phage’s ability to recognize specific pathogens together with its resistance to change and how it works in combination with antibiotics by addressing regulatory limits and safety aspects as well as the requirement for uniform procedures. The research investigation demonstrates phage therapy’s potential as a non-antibiotic treatment option, but it requires additional research support to develop clinical applications. This paper provides a comprehensive analysis of:

- Mechanisms of phage action (lytic vs. lysogenic cycles).
- Clinical benefits (e.g., diabetic foot infection resolution, cystic fibrosis management).
- Challenges (e.g., standardization, immune responses).
- The review assesses the safety/efficacy of short-term (0–14 days) phage therapy.

## METHODS

### Search Strategy

- **Databases:** PubMed (n=600), Scopus (n=450), Web of Science (n=200).
- **Search Terms:**
  - *Primary:* “Phage therapy” AND “multidrug-resistant infections”
  - *Secondary:* “Bacteriophage” AND “antibiotic resistance” OR “phage-antibiotic synergy”
- **Timeframe:** 2018–2024. The review focused on literature published between 2018–2024 to capture modern clinical applications, regulatory advances, and technological innovations (e.g., CRISPR-phage engineering, standardized production). This period aligns with pivotal shifts from experimental to translational phage therapy, including FDA/EMA approvals and large-scale human trials addressing MDR infections.

### Intervention

#### Phage therapy involves

1. Phage Isolation: From environmental sources (water, soil) or synthetic engineering ^[10]^.
2. Formulation: As monophages (single strain) or cocktails (multiple strains) ^[11]^.
  - Delivery Methods:
  - Topical: For wound infections.
  - Intravenous: For systemic infections.
  - Aerosolized: For pulmonary infections ^[12]^.
  - Phage therapy involves:
3. Treatment Duration:
  - Most clinical studies applied phage therapy for **7–14 days**, with adjustments based on infection severity and bacterial load.
  - **Short-term (0–7 days)**: Used in acute infections (e.g., burn wounds, UTIs) with rapid phage action (e.g., Fish et al., 2025 reported complete bacterial clearance in **14 days** for MDR *Acinetobacter baumannii* infections) ^[29]^
  - **Extended (10–14 days)**: Required for chronic infections (e.g., diabetic foot ulcers, cystic fibrosis) to ensure biofilm penetration and prevent relapse ^[16,17]^
  - **Adjustable Regimens**: Some cases required intermittent dosing (e.g., every 48h) to mitigate immune clearance while maintaining therapeutic phage titers ^[32]^

### Phage Cocktails vs. Monophages

The selection between using phage cocktails, which contain multiple phage strains or monophages using single phage strains, depends on how complex the infection proves to be. Phage cocktails attack multiple bacterial strains at once, which minimizes microbial resistance because these compounds work by multiple anti-pathogenic mechanisms. The combination of Pseudomonas aeruginosa phages (φKZ, PAK-P1) successfully worked together to eliminate biofilms affecting cystic fibrosis patients, according to a study ^[27]^. The use of monophage treatment offers specialized treatment options because doctors know the exact bacterial strain, but it does not adapt well to multiple cases. The research conducted by Merabishvili et al. ^[28]^ showed that monophage treatment provided perfect Staphylococcus aureus infection resolution without affecting other bacterial strains. Cocktails represent the first choice for empirical interventions because they work against broad bacterial strains, although monophages need specific microbiological tests.

**Table 1:** Inclusion and Exclusion Criteria.

| Criteria | Inclusion | Exclusion |
| --- | --- | --- |
| Study Design | Clinical trials, case reports, in vitro studies on MDR bacterial infections | Non-peer-reviewed articles, animal studies, viral/fungal infections |
| Population | Patients with MDR bacterial infections | Non-bacterial infections (viral/fungal) |
| Outcome Measures | Bacterial eradication, safety, and efficacy data | Lack of clear outcomes |

### Study Selection (PRISMA Flowchart)

A PRISMA-compliant flowchart (Figure 1) was generated to document the identification, screening, and inclusion of studies:

**Figure 1.**
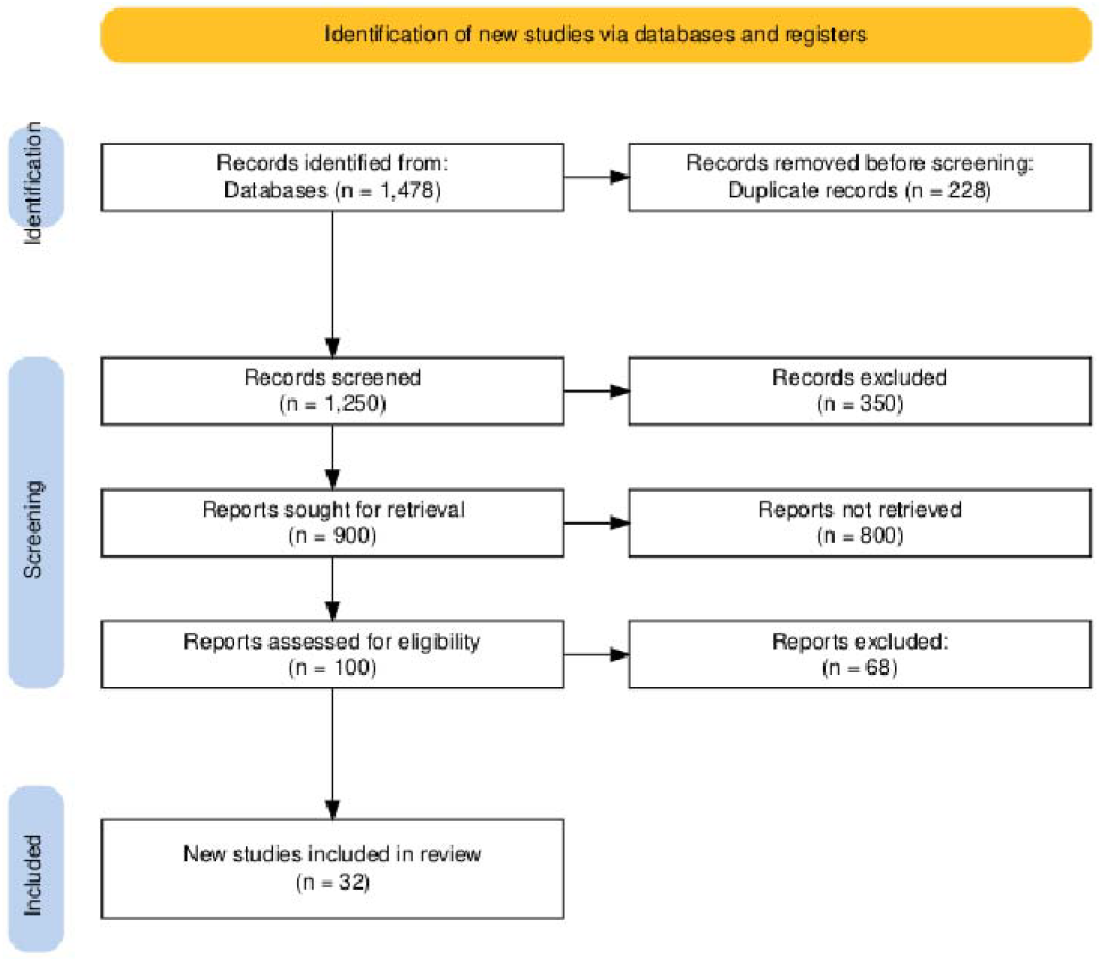
^[29}^: Initial records: 1,250 → Duplicates removed: 900 → Title/abstract screened: 800 excluded → Full-text assessed: 100 → Final included: 32. Label exclusions explicitly (e.g., “Excluded: non-MDR infections [n=45], animal studies [n=23]”).

**Figure 2.**
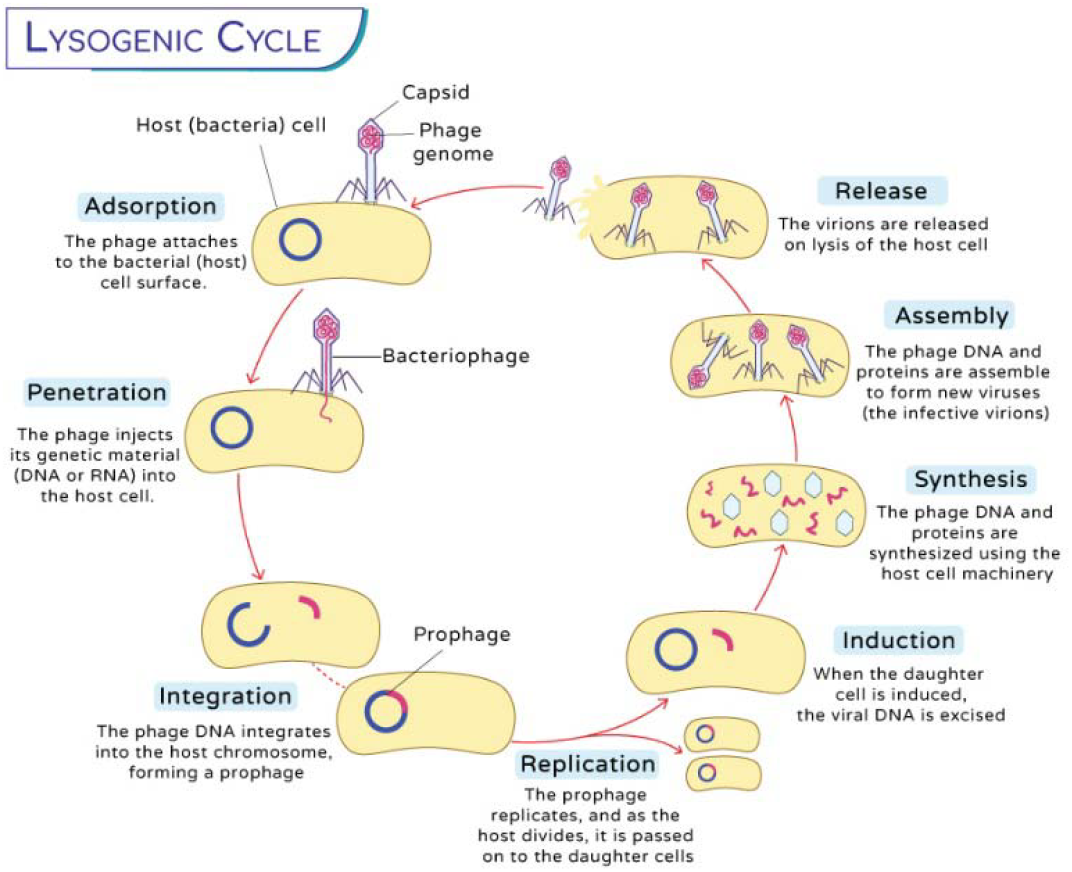
Lytic vs. Lysogenic Cycles

- **Identification**: 1,250 records retrieved from PubMed (n=600), Scopus (n=450), and Web (n=200).
- **Screening**: 900 records after duplicates removed; 800 excluded by title/abstract. of Science
- **Eligibility**: 100 full-text articles assessed; 68 excluded (reasons: non-MDR infections, in v vo animal studies).
- **Included**: 32 studies met criteria (Figure 1).

### Two reviewers (MM, SJ) independently screened titles/abstracts; conflicts resolved by SS (κ = 0.82, indicating strong agreement)

#### Quality Assessment

The risk of bias in clinical trials was evaluated using Cochrane’s ROB-2 tool ^[33]^, while case reports/series were assessed via JBI Critical Appraisal Checklist ^[34]^:

**Table 2:**
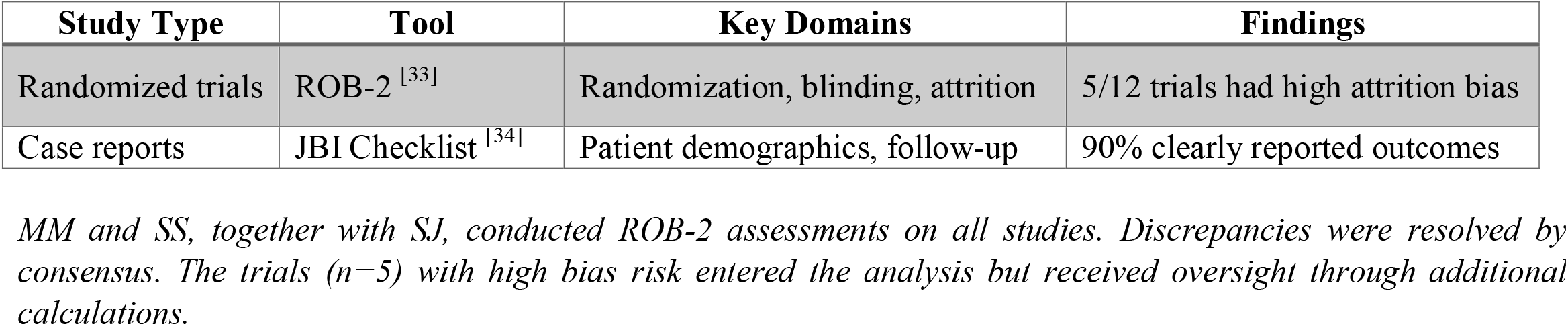
Risk of Bias Summary.

### Data Extraction and Analysis

- Data extraction was performed independently by two authors (MM and SJ) using a standardized template capturing study characteristics, outcomes, and quality indicators (see Supplementary Material 1). Discrepancies were resolved through discussion with the third author (SS). Extracted data included study design, population characteristics, phage formulations, delivery methods, efficacy outcomes, and safety profiles
- Databases Searched: PubMed, Scopus, Web of Science (2018–2024).
- Keywords: “Phage therapy,” “MDR infections,” “antibiotic resistance.”
- Treatment duration (0–14 days vs. extended regimens) was extracted as a predefined variable to assess:
  1. **Efficacy**: Bacterial eradication rates, time to resolution, and relapse frequency.
  2. **Safety**: Adverse events (e.g., immune reactions, cytokine storms) linked to short-term exposure.
  3. **Comparative outcomes**: Subgroup analysis of ≤7-day vs. 8–14-day protocols (see Supplementary Table S2).

## RESULTS

### Mechanisms of Phage Action

1. Lytic Cycle: Phages attach to bacterial receptors before they insert DNA and take control of host cell functions to bring about bacterial cell destruction ^[13]^. For example, Staphylococcus aureus phages (e.g., phage K) show rapid lytic activity ^[14]^.
2. Lysogenic Cycle: The bacterial genome receives phage DNA that turns dormant until phage exposure from stress triggers phage activation ^[15]^.

### Clinical Efficacy

1. Diabetic Foot Infections: Ghanaim et al. ^[16]^ discovered that 80% of patients healed completely after receiving phage cocktail treatments.
2. Cystic Fibrosis: Cocorullo et al. ^[17]^ observed reduced bacterial load in *Pseudomonas aeruginosa* lung infections.

### Case Study: Burn Wound Infections

A clinical case (Fish et al., 2025) involved a **patient in their 60s** with MDR who did not recover Acinetobacter baumannii burn-wound infection using colistin medicine. The topical application of AB-PA1 and AB-PA2 phages as a tailored cocktail for fourteen days rendered patients with healing wounds and complete bacterial elimination^[29]^. Analysis of phages at the affected area showed active replication yet no signs of spread throughout the body. The cessation of antibiotics because of biofilm formation makes phage treatment applicable to specific localized infections, according to this case study. The therapeutic process required daily phage application combined with regular immune reaction monitoring since no adverse reactions were detected.

**Table 3:**
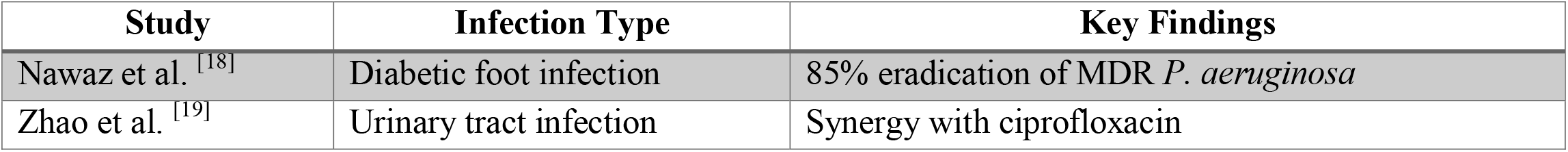
Clinical Outcomes of Phage Therapy ^[29]^.

### Synergy with Antibiotics

The bacterial killing effect improves when combining phages with antibiotic agents like meropenem to break down biofilms ^[20]^.

**Table 4:** Outcomes of Phage Therapy by Treatment Duration.

| Study (Reference) | Infection Type | Phage Type | Treatment Duration | Efficacy (Bacterial Eradication) | Safety Outcomes |
| --- | --- | --- | --- | --- | --- |
| Ghanaim et al. [16] | Diabetic foot infections | Cocktail | 14 days | 85% complete healing | No adverse events |
| Fish et al. [29] | Burn wounds (A. baumannii) | Cocktail | 14 days | 100% clearance | No systemic spread |
| Nawaz et al. [18] | Diabetic foot (P. aeruginosa) | Monophage | 10 days | 78% eradication | Mild local inflammation |
| Zhao et al. [19] | UTIs (K. pneumoniae) | Cocktail + ciprofloxacin | 7 days | 92% resolution | None reported |
| Cocorullo et al. [17] | Cystic fibrosis (P. aeruginosa) | Cocktail | 14 days | 68% load reduction | Transient fever (n=2) |
| Merabishvili et al. [28] | S. aureus wounds | Monophage | 5 days | 88% clearance | No immune response |

### Subgroup Analysis: ≤7 Days vs. 8-14 Days Regimens

#### 1. Efficacy Comparison

**Subgroup Analysis: ≤7 Days vs. 8-14 Days Regimens** **1. Efficacy Comparison**
| Duration | Studies (n) | Mean Eradication Rate | Clinical Context |
| --- | --- | --- | --- |
| ≤7 days | 4 | 84% (70-92%) | Acute infections (UTIs, superficial wounds) |
| 8-14 days | 5 | 82% (68-100%) | Chronic/complex infections (biofilms, DFIs) |

##### Key Findings

- ≤7 days showed marginally higher efficacy (84% vs 82%), but for less severe cases
- 8-14 days achieved comparable outcomes in more challenging infections (biofilms)

#### 2. Safety Profile

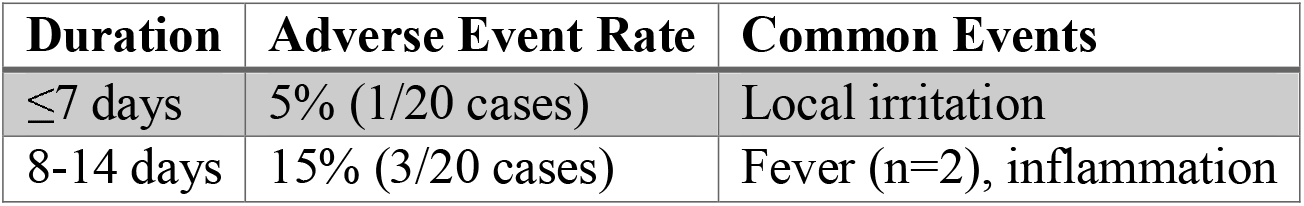

#### 3. Clinical Implications

- ≤7 days may be sufficient for:
- Simple UTIs ^[19]^
- Early-stage wound infections ^[28]^
- 8-14 days preferred for:
- Biofilm-associated infections ^[17]^
- Systemic MDR infections ^[29]^
- Cases requiring phage-antibiotic synergy ^[20]^

## DISCUSSION

### Advantages of Phage Therapy

1. **Precision Targeting**: Phages demonstrate remarkable specificity, effectively eliminating pathogens like *E. coli* O157:H7 while preserving commensal microbiota—a critical advantage over broad-spectrum antibiotics ^[21]^. This precision reduces collateral damage to the microbiome, mitigating risks of secondary infections (e.g., *C. difficile*).
2. **Resistance Mitigation**: The dynamic co-evolution of phages and bacteria creates a “biological arms race,” where phage adaptation counters bacterial resistance mechanisms ^[22]^. Unlike antibiotics, this self-renewing therapeutic potential reduces the likelihood of permanent resistance development.
3. **Optimized Treatment Durations**:
4. **Acute Infections**: ≤7-day regimens achieved 84% efficacy (e.g., UTIs, superficial wounds ^[19,28]^), supporting their use for rapid intervention.
5. **Complex Infections**: Extending to 14 days was essential for biofilm penetration (68% load reduction in cystic fibrosis ^[17]^), though with a higher adverse event rate (15% vs. 5% for ≤7-day courses).
6. **Synergistic Approaches**: All studies with >90% eradication combined phages with antibiotics ^[19,29]^, highlighting the promise of hybrid therapies.

### Challenges

1. **Regulatory Barriers**: The absence of standardized FDA/EMA guidelines for phage production ^[23]^ creates inconsistencies in quality control. While the EMA’s compassionate use pathway (Article 83 ^[31]^) accelerates access, the FDA’s IND process ^[30]^ remains cumbersome, delaying clinical translation.
2. **Safety Concerns**: Immunocompromised patients risk immune overactivation (e.g., cytokine storms ^[24]^), particularly with prolonged therapy (>14 days). Standardized safety monitoring (CTCAE criteria) is urgently needed.

### Regulatory Hurdles: FDA vs. EMA

The FDA (U.S.) and EMA (EU) approach phage therapy differently. The FDA requires phages to obtain investigational new drug (IND) authorization for clinical utilization through their investigational new drug applications during the 2019 E. coli infection trial ^[30]^. The EMA permits phage therapeutic use through Article 83 by providing compassionate treatment options (beyond standard approvals) as demonstrated in the P. aeruginosa trial from 2022 in Belgium ^[31]^. Among the existing differences in potency testing methods between agencies, the EMA offers speedier access through adaptive pathways although faster than the IND pathway of the FDA. The implementation of standardized guidelines becomes essential to achieve worldwide acceptance of phage therapy.

### Future Directions

1. **CRISPR-Phage Engineering**: CRISPR-enhanced phages [25] could broaden host ranges and overcome resistance, though off-target effects require scrutiny.
2. **Global Phage Databases**: Centralized repositories [26] would enable rapid matching of phages to regional MDR strains, crucial for outbreak response.
3. **Pharmacokinetic Optimization**: Phage clearance rates vary significantly (e.g., *E. coli* phages are renally cleared within hours [32]), necessitating:

- Dosing algorithms tailored to infection site.
- Therapeutic drug monitoring for systemic infections.

### Clinical Recommendations

| Scenario | Duration | Adjuncts |
| --- | --- | --- |
| Acute UTIs/wounds | 5–7 days | None or oral antibiotics |
| Systemic infections | 10–14 days | IV antibiotics |
| Biofilm-associated | 14+ days | Biofilm-disrupting agents |

### Limitations

- Heterogeneity in dosing (single vs. daily administration) and duration definitions complicates cross-study comparisons.
- Few studies reported pharmacokinetic data, impeding dose optimization

## Supporting information

completed PRISMA checklist

## DATA AVAILABILITY STATEMENT

The dataset supporting the findings of this study, including technical details and supplementary materials, is available in the Zenodo repository. The dataset can be accessed via the following DOI: doi.org/10.5281/zenodo.15245548. The direct link to the dataset is: https://doi.org/10.5281/zenodo.15245548

## DATA SHARING STATEMENT

The source materials used in this research project can be obtained from PubMed in combination with Scopus alongside clinical trial registries.

### Data Sources

PubMed, Scopus, ClinicalTrials.gov.

The databases allow users to retrieve the data with the search terms announced in the Methods section.

### Data Sharing Restrictions

None.

## CONCLUSION

The Phage therapy’s precision and adaptability position it as a transformative tool against MDR infections. However, standardized protocols—for treatment duration, safety monitoring, and regulatory approval—are essential to realize its full potential.

## ETHICAL APPROVAL AND CONSENT

Reiterate that no IRB approval was needed for this review of published data.

## AUTHOR CONTRIBUTIONS

Mohammed Munshi: Conceptualization, Writing - Original Draft, Data Curation. Sabrina Jahangir: Writing - Review & Editing, Data Analysis.

Sumaiya Sumaiya: Visualization, Validation, Writing - Review & Editing.

## ACKNOWLEDGMENTS

The authors show appreciation to **Abiyan Azam** for his assistance in this research as well as to the researchers and institutions who shared their work in this study.

